# A model-based approach to improve intranasal sprays for respiratory viral infections

**DOI:** 10.1101/2022.01.26.22269854

**Authors:** Saikat Basu, Mohammad Mehedi Hasan Akash, Yueying Lao, Pallavi A Balivada, Phoebe Ato, Nogaye K Ka, Austin Mituniewicz, Zachary Silfen, Julie Suman, Arijit Chakravarty, Diane Joseph-McCarthy

## Abstract

Drug delivery for viral respiratory infections, such as SARS-CoV-2, can be enhanced significantly by targeting the nasopharynx, which is the dominant initial infection site in the upper airway, for example by nasal sprays. However, under the standard recommended spray usage protocol (“Current Use”, or CU), the nozzle enters the nose almost vertically, resulting in sub-optimal deposition of drug droplets at the nasopharynx. Using computational fluid dynamics simulations in two anatomic nasal geometries, along with experimental validation of the generic findings in a different third subject, we have identified a new “Improved Use” (or, IU) spray protocol. It entails pointing the spray bottle at a shallower angle (almost horizontally), aiming slightly toward the cheeks. We have simulated the performance of this protocol for conically injected spray droplet sizes of 1 – 24 *μ*m, at two breathing rates: 15 and 30 L/min. The lower flowrate corresponds to resting breathing and follows a viscous-laminar model; the higher rate, standing in for moderate breathing conditions, is turbulent and is tracked via Large Eddy Simulation. The results show that (a) droplets sized between ∼ 6 – 14 *μ*m are most efficient at direct landing over the nasopharyngeal viral infection hot-spot; and (b) targeted drug delivery via IU outperforms CU by approximately 2 orders-of-magnitude, under the two tested inhalation conditions. Also quite importantly, the improved delivery strategy, facilitated by the IU protocol, is found to be robust to small perturbations in spray direction, underlining the practical utility of this simple change in nasal spray administration protocol.

## 1. Introduction

The global respiratory pandemic^1^ caused by the severe acute respiratory syndrome coronavirus 2 (SARS-CoV-2) has thrust the field of fluid mechanics back into public eye, perhaps for the first time since the era of 1960s’ space race.^2^ Flow physics plays an essential role in almost every aspect of respiratory viral infections; none the more so than in targeted delivery of drugs to the infection hot-spots along the airway. Upper airway sites, in specific the ciliated epithelial cells that line the back of the nasal cavity at the nasopharynx (see Fig. 1) and are rich in angiotensin-converting enzyme 2 (ACE2) surface receptors, have been marked out^3–5^ as the trigger zones for infection onset owing to SARS-like airborne viral respiratory pathogens. An early intervention method that can target the initial dominant infection site, i.e. the nasopharynx, is hence imperative for limiting asymptomatic transmission of the exhaled pathogenic particulates as well as for preventing systemic lower airway progression of the disease in a host, aggravating toward severe illness.^6, 7^ Of critical interest here, based on the brisk pace at which lower airway infections ensue after the emergence of initial symptoms, it has been conjectured^3, 8, 9^ that the nasopharynx also acts as the seeding zone for spread of the disease to the lungs via lower airway aspiration of virus-laden boluses of nasopharyngeal fluids. Another concern is the mutation rate of SARS-CoV-2 and how the nature of the fitness landscape renders the virus amenable to evolving, potentially resulting in more virulent strains.^10^ A nasal spray – that can administer nasal hygiene products, prophylactics, and antiviral agents – would address these concerns if it can efficiently deliver the pharmaceutics at the virus-affected upper airway sites, thereby reducing the risk of viral droplet/aerosol shedding as well as mutation within the host.

**Figure 1:**
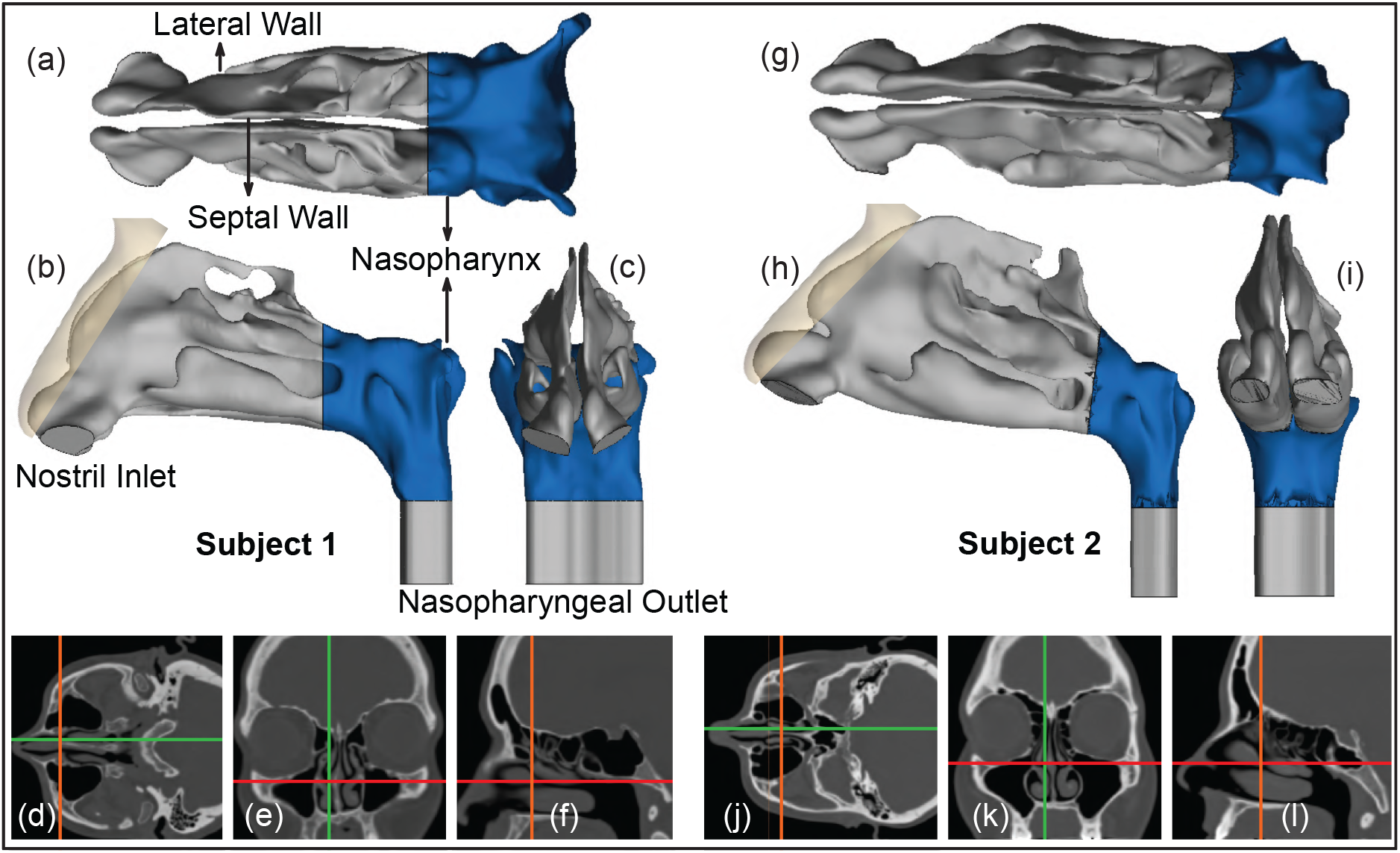
Panels (a) – (c) respectively show the axial, sagittal, and coronal views of the computed tomography (CT) based upper airway reconstruction in Subject 1. Panels (d) – (f) depict representative CT slices for the same subject. Therein, the green lines in (d) and (e) correspond to the location of the sagittal section shown in (f); the orange lines in (d) and (f) correspond to the location of the coronal section shown in (e); the red lines in (e) and (f) correspond to the location of the axial section shown in (d). Panels (g) – (i) respectively show the axial, sagittal, and coronal views of the CT-based upper airway reconstruction in Subject 2. Panels (j) – (l) depict representative CT slices for the same subject. Therein, the green lines in (j) and (k) correspond to the location of the sagittal section shown in (l); the orange lines in (j) and (l) correspond to the location of the coronal section shown in (k); the red lines in (k) and (l) correspond to the location of the axial section shown in (j). The nasopharynx has been marked in blue in panels (a) – (c) and (g) – (i).

While the nasal sprays do provide a simple, yet robust, drug delivery modality, especially during the infection onset phase of respiratory viruses; the choice still comes with at least two key open questions, *viz*. (a) what are the intranasally sprayed drug droplet sizes that would maximize targeted delivery at the initial dominant infection site, the nasopharynx?; and (b) is there a way to revise the nasal spray usage protocols, to enhance the delivery of drugs at the infected site?

This study addresses the above questions through implementing experimentally-validated computational fluid dynamics (CFD) modeling of the respiratory transport process in computed tomography (CT)-based anatomically realistic upper airway geometries. The related simulations replicate sprayed drug transmission against two different ambient inhalation rates, *viz*. 15 and 30 L/min; standing in respectively for relaxed and moderate steady breathing conditions.^11^ Preliminary findings pertaining to this work have been presented at the American Physical Society’s Division of Fluid Dynamics Annual Meeting 2021.^12^

## 2. Materials and methods

### 2.1. Anatomic upper airway reconstruction

The *in silico* upper airway geometries used here were reconstructed from the de-identified medical-grade CT imaging data derived from two healthy test subjects. Subject 1 was a Caucasian female in the age range 61-65 years; Subject 2 was a Caucasian female in the age range 36-40 years. Use of the archived and anonymized medical records was approved with exempt status by the Institutional Review Board (IRB) for the University of North Carolina (UNC) at Chapel Hill, with informed consent waived for retrospective use in computational research.

In terms of imaging resolution, the CT slices of the airway cavities were extracted at coronal depth increments of 0.348 mm in Subject 1’s scans and 0.391 mm in Subject 2’s scans. Digitization of the anatomic airspaces was carried out on the image processing software Mimics Research v18.0 (Materialise, Plymouth, Michigan), using a radio-density delineation range of -1024 to -300 Hounsfield units, and was complemented by clinically-monitored hand-editing of the selected pixels to ensure anatomic accuracy. The output STL (stereolithography) geometries were then spatially meshed on ICEM-CFD 2019 R3 (ANSYS Inc., Canonsburg, Pennsylvania) with minute volume elements. Therein to confirm grid-independent solutions, established mesh-refinement protocols^13, 14^ were followed such that each computational grid contained more than 4 million unstructured, graded, tetrahedral elements. To enable accurate tracking near tissue surfaces, further mesh refinement involved adding three prism layers at the cavity walls, with 0.1-mm thickness and a height ratio of 1.

### 2.2. Simulation of breathing transport and drug delivery

Inhalation parameters for gentle-to-moderate breathing conditions were numerically replicated at 15 and 30 L/min.^11^ The lower flow rate commensurate with resting breathing is dominated by viscous-laminar steady-state flow physics.^15–20^ The higher flow rates however trigger shear-induced^21–23^ flow separation from the tortuous cavity walls, resulting in turbulence,^24–27^ which was tracked through Large Eddy Simulation (LES), with sub-grid scale Kinetic Energy Transport Model^28^ accounting for the small-scale fluctuations. The computational scheme on ANSYS Fluent 2019 R3 employed a segregated solver, with SIMPLEC pressure-velocity coupling and second-order upwind spatial discretization. Solution convergence was monitored by minimizing mass continuity and velocity component residuals, and through stabilizing mass flow rate and static pressure at airflow outlets (see the nasopharyngeal outlet location in Fig. 1). For the pressure gradient-driven laminar airflow solutions, the typical execution time for 5000 iterations was 2–3 hours with 4-processor based parallel computations operating at 3.1 GHz speed on Xeon nodes. Additionally, the LES computations each required a run-time of 1–2 days, for a pressure-driven simulated flow interval of 0.25 s, with a time-step of 0.0001 s. To realistically capture the inhaled warmed-up air transport along the respiratory pathway, its density and dynamic viscosity were set at 1.204 kg/m^3^ and 1.825 *×* 10^−5^ kg/m.s, respectively.

Spray dynamics against the ambient airflow was tracked via Lagrangian-based inert discrete phase simulations with a Runge-Kutta solver, with localized droplet clustering along intranasal tissues obtained through numerically integrating the transport equations that consider airflow drag, gravity, and other body forces relevant for small particulates, e.g., the Saffman lift force, and by implementing a no-slip trap boundary condition on the cavity walls. Note that Brownian effects were neglected in view of the tracked droplet sizes. The drug formulation density was set to 1.5 g/ml. All simulations released monodispersed inert drug droplets ranging in diameters from 1 – 24 *μ*m, with 3000 monodispersed inert droplets being released during each iteration. The droplets were injected into the airspace from a single source point where the spray nozzle is located, streaming out in a hollow-cone shape, mimicking the action of a nasal spray; this method of release is referred to as a cone injection. The Valois VP7, an affordable mass-produced pharmaceutical nasal spray pump, with its accompanying dimension properties, such as plume angle and initial spray velocity, was used as an initial point of reference for the cone injections.^29^ The droplets were given an initial velocity of 10 m/s^30^ and a total flow rate of 1*×*10^−20^ kg/s. The plume angle and insertion depth were selected^15^ to be 27.93^°^ and 5 mm, respectively; by varying the spray direction – an optimal usage condition that augments droplet deposition at the target site was identified. See our earlier publications^5, 15^ for additional details on the numerical setup.

### 2.3. On how to hold the spray bottle

A key parameter for targeted delivery is the direction of the nasal spray axis, as the sprayed droplet trajectories are often inertia-dominated.^15, 18, 31, 32^ Instructional ambiguities^33, 34^ point toward a lack of definitive knowledge on the best ways to use a nasal spray device, with different commercial sprayers often offering somewhat contrasting recommendations. There is, however, a consensus that the patient should tilt her/his head slightly forward, while holding the spray bottle upright.^33, 35^ There is an additional clinical recommendation to avoid pointing the spray directly at the septum, which is the separating cartilaginous wall between the two sides of the nasal cavity. These suggestions were adopted in our standardization^15, 36^ of “Current Use” (CU) protocol for topical sprays. The digital models were inclined forward by an angle of 22.5^°^, and the vertically-placed upright^33^ spray axis was aligned closer to the lateral nasal wall, at one-third of the distance between the lateral side and septal wall. Finally, the spray bottle was placed at the nostril to penetrate 5-mm into the airspace, to conform with the package recommendations of commercial sprayers^35^ for a “shallow” intranasal nozzle placement.

While the CU protocol would provide the acceptable state-of-art for targeted drug delivery with nasal sprayers, the key focus of this study was to perturb that spray direction to test alternate protocols that bear the promise to improve delivery of drugs at the nasopharyngeal infection site. Our earlier findings^15^ showed that to target the clinical site of ostiomeatal complex, or OMC (a key target site for corticosteroid-based topical therapeutic management for chronic rhinosinusitis^15, 19^ and allergic rhinitis^37^), the spray axis should be oriented to pass through the OMC itself. The inertial motion of the sprayed particulates assists such transport mechanism. Taking cue, to optimize the spray administration protocol in the current study, we oriented the nozzle such that the spray axis passes through the nasopharynx, and have named the strategy as “Improved Use”, or IU protocol. When determining the IU direction, it was important to satisfy three conditions as a way of ensuring the optimal placement of a nasal spray for drug release: (i) the extended spray axis for the IU protocol must intersect the nasopharynx; (ii) the spray axis must not cut through the septal wall; and (iii) the axis should intersect the lateral wall in the posterior part of the nasal cavity. See the cartoonized Fig. 2 for a broad-spectrum visual difference between the presently recommended CU and the to-be-tested IU protocols. Additionally, Figs. 3 and 4 depict the spatial distinctions in spray placement between the IU and CU protocols, in the two test subjects, as visible from the sagittal perspective.

**Figure 2:**
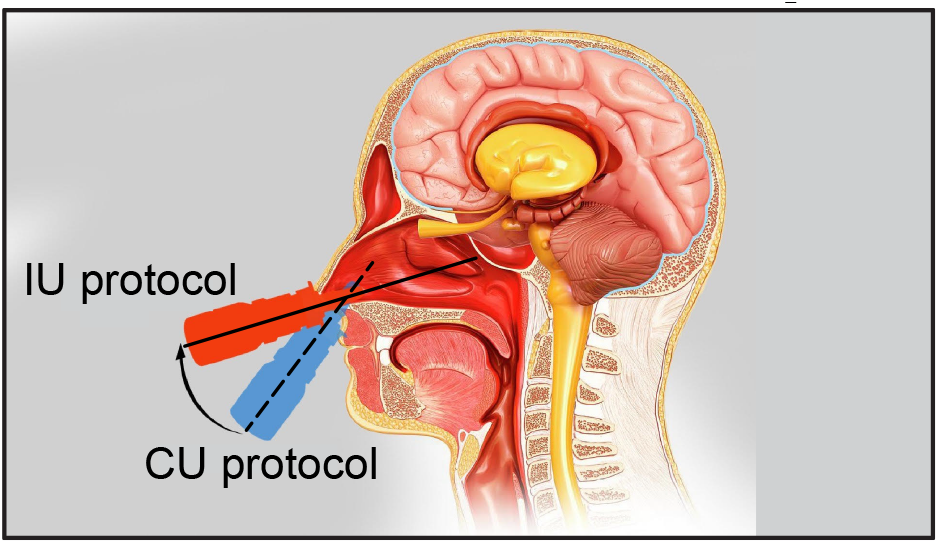
The schematic shows the two tested nasal spray usage protocols, *viz*. “Current Use” (or CU, represented by the dashed line) and “Improved Use” (or IU, represented by the solid line). Cartoon illustration is by the Dr. Ferrer Biopharma (Hallandale Beach, FL) graphics design team.

**Figure 3:**
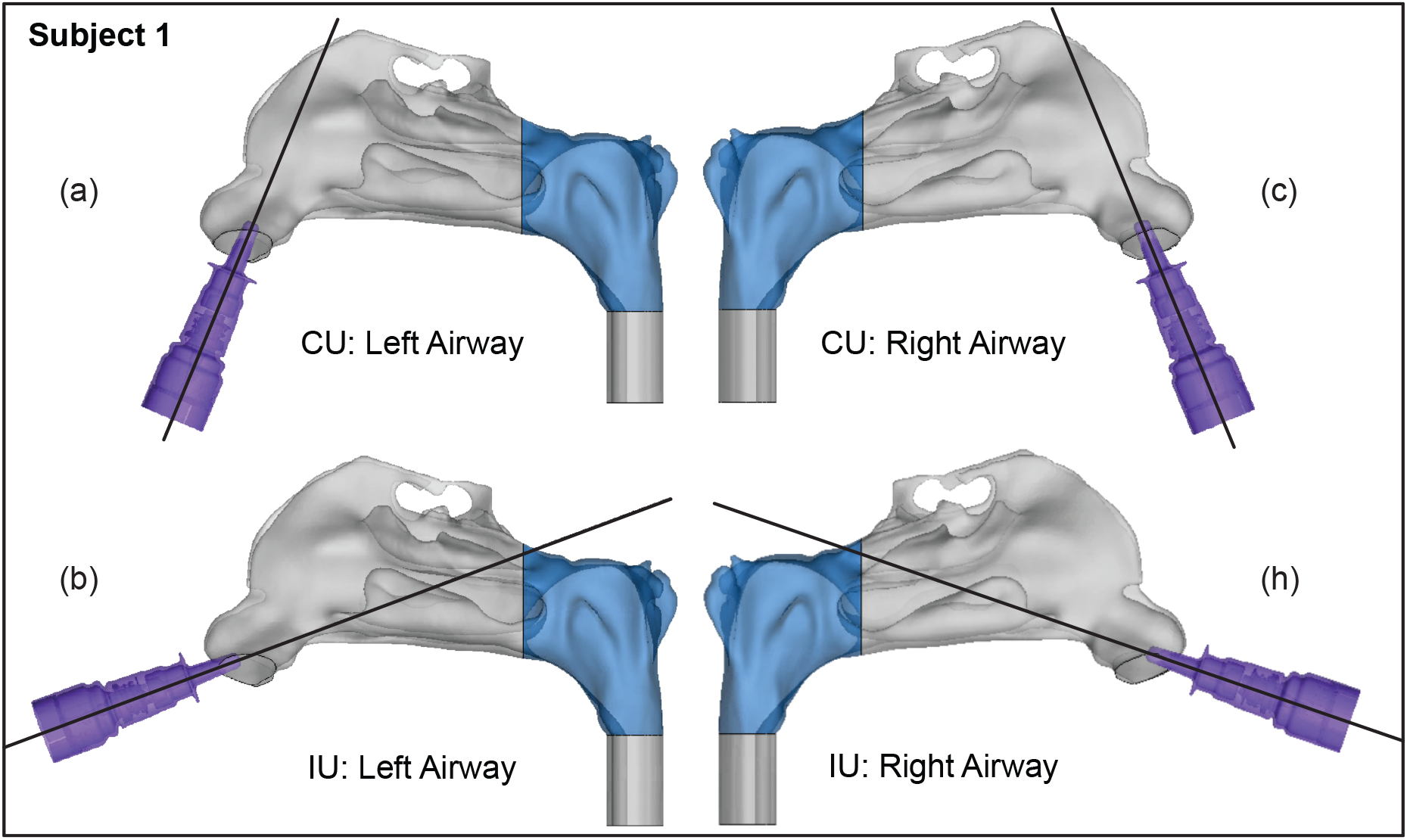
Spatial differences between the Current Use (CU) and Improved Use (IU) spray placement protocols, as visible sagittally in Subject 1. Nasopharynx is marked in blue.

**Figure 4:**
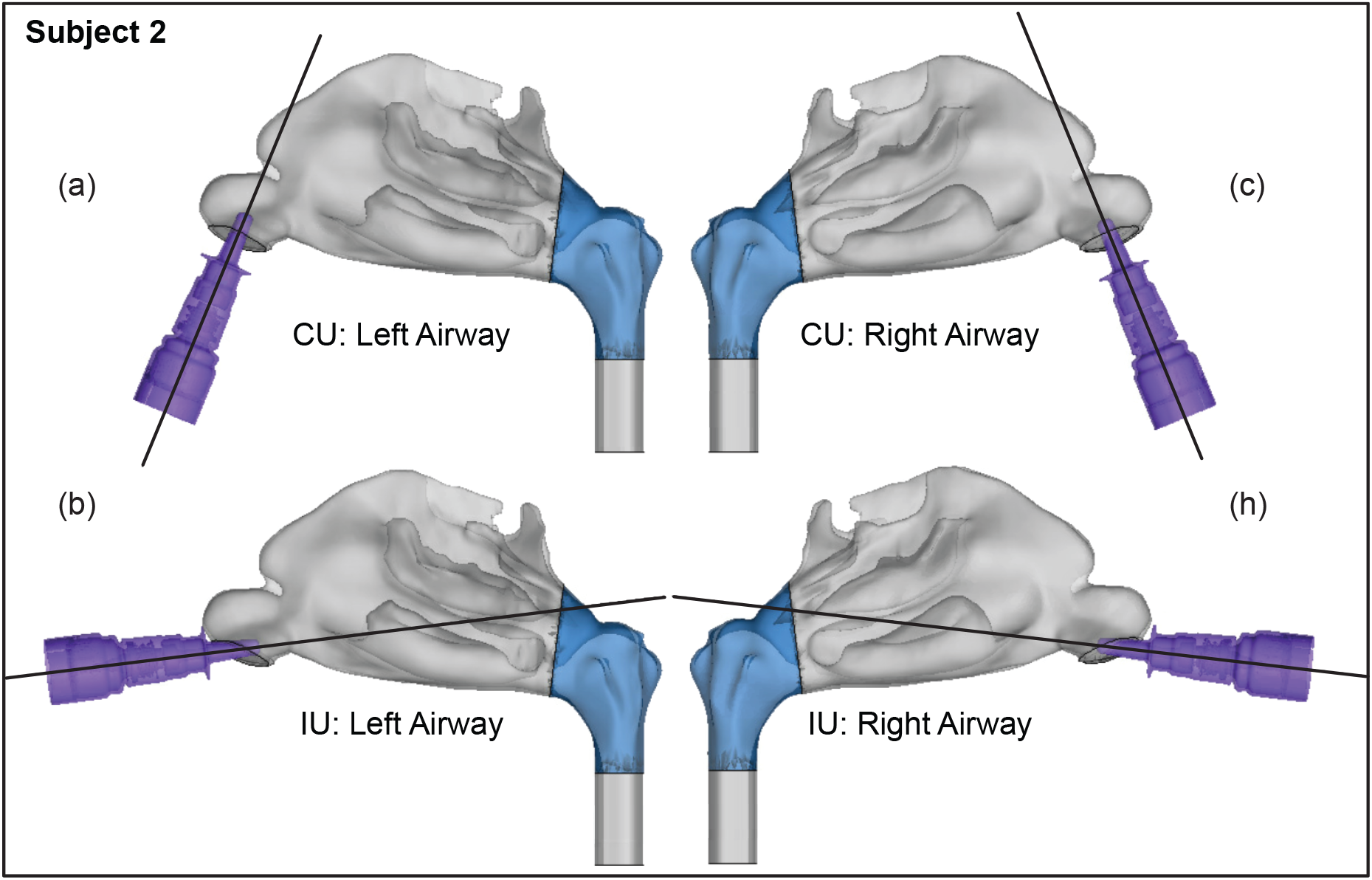
Spatial differences between the Current Use (CU) and Improved Use (IU) spray placement protocols, as visible sagittally in Subject 2. Nasopharynx is marked in blue.

### 2.4. Tolerance sensitivity analysis

Once the IU for an airway reconstruction was determined (following guidelines described in Section 2.3), a tolerance sensitivity study was performed to assess how far the user could deviate from the determined IU spray direction and still get similar regional drug deposition results, or in other words how robust (or, on the contrary, user-sensitive) the chosen IU direction really is.

To generate the new perturbed axes in the *in silico* space, a 1-mm radius circle was created perpendicular to the perturbed direction either 5-mm or 10-mm away from the central point on the nostril plane of each model. The two different distances were chosen in order to test the sensitivity of the results to different perturbation trends. The 5-mm method was performed on the left nostril of the subjects, while the 10-mm method was performed on the right nostril. Five peripheral points equidistant from each other were then selected on the circle created. The axis formed between the centroid point on the nostril plane and the peripheral point on the circle determined the new perturbed direction. In all, five additional perturbed spray axes were created, henceforth referred to as PD 1 – 5. For each new perturbed direction, the injection point was selected by measuring 5-mm from the centroid on the nostril plane, toward the nasopharynx. This was performed for both the left and right nostrils of Subjects 1 and 2. Each new identified PD axis was evaluated using the criteria developed to identify the IU direction, and drug delivery simulations were performed following the methods described in Section 2.2. The results of the tolerance simulations were analyzed for congruity using Pearson’s correlation coefficient.

### 2.5. Experimental validation of computationally predicted spray performance

To extract a sense of real spray performance that could be projected from the *in silico* framework, we linked the computationally predicted nasopharyngeal droplet deposition efficiencies with the size distribution of droplets (see Fig. 5) in two actual over-the-counter spray products: Flonase™ (Fluticasone Propionate) and Nasacort™ (Triamcinolone Acetonide). Both are commonly prescribed medications and are commercially available. Four units of each product were tested at Next Breath, an Aptar Pharma company (Baltimore, MD, USA). The team measured the plume geometry through a SprayVIEW® NOSP, which is a non-impaction laser sheet-based instrument. With the droplet sizes in a spray shot following a log-normal distribution, the droplet size distribution (where droplet diameters are represented by *x*) can be framed as a probability density function:^38^

**Figure 5:**
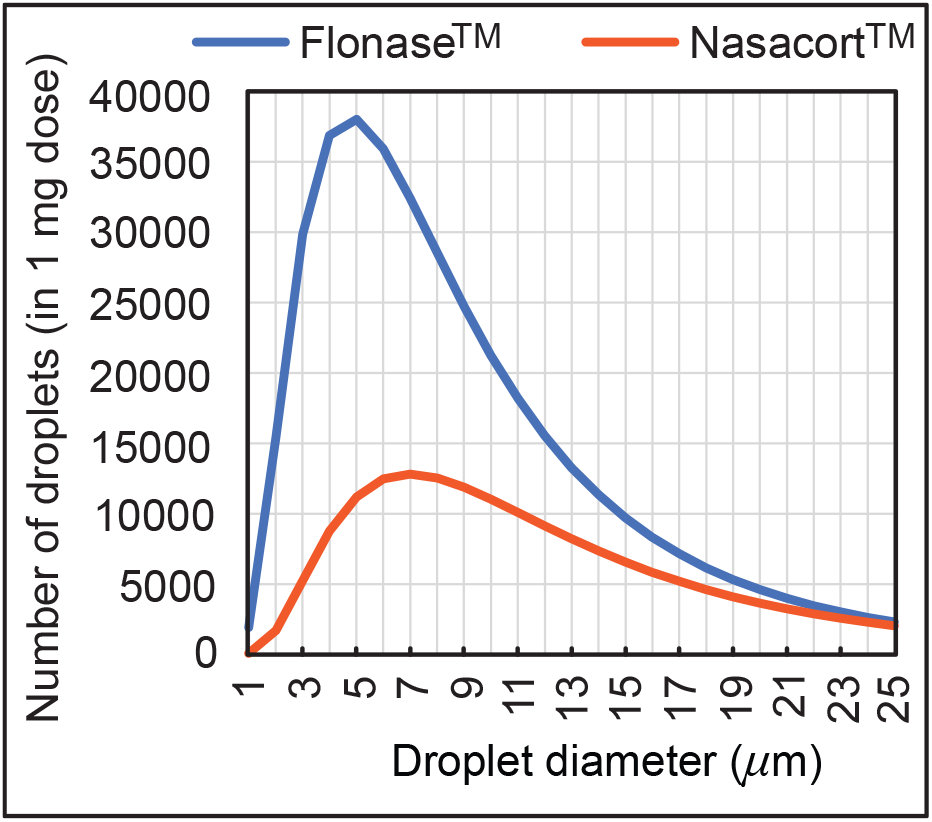
Measured distribution of droplet sizes in 1-mg sprayed mass from over-the-counter Flonase™(Fluticasone Propionate) and Nasacort™ (Triamcinolone Acetonide) spray products, over the test size range of ∼ 1 – 24 *μ*m used for *in silico* tracking. Note that rigorous testing for droplets > 24 *μ*m clearly show^5^ that they would deposit along the anterior nasal cavity and will not directly land at the posterior target site of nasopharynx.

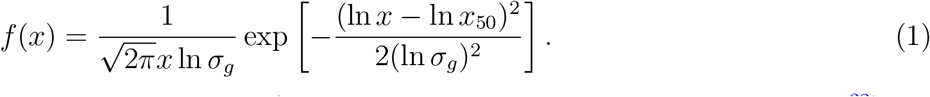

Here the mass median diameters (alternatively, the geometric mean diameter^32^) for Flonase™ and Nasacort™ were respectively, *x*_50_ = 37.16 *μ*m and 43.81 *μ*m; the corresponding geometric standard deviations were respectively, *σ*_*g*_ = 2.080 and 1.994. The latter quantifies the span of the droplet size data. Note that the measurements were also collected with and without a saline additive in the sprayer, with the tests returning similar droplet size distributions. The reader is referred to our previous publications^15, 18^ for additional details.

To test the validity and extensibility of the computational predictions derived for real sprays, we subsequently performed multiple runs of physical spray experiments with 10-ml boluses (for measurable posterior deposits) of watery solutions injected through a 3D-printed anatomically realistic airway cavity of a different subject, Subject 3 (a Caucasian male belonging to the age range 41-45 years; use of the subject’s de-identified imaging data with CT-slice resolution of 0.352 mm was approved with exempt status by the UNC Chapel Hill IRB for retrospective use). Printing of the related anterior soft plastic part on a Connex3™ 3D printer was carried out using polymer ink-jetting process on Tangogray FLX950 material, approximately mimicking the material properties of the external nares and the internal tissues and cartilages. The 3D-printed cavity extent terminated just before the nasopharynx, thereby allowing us to measure the outflow volume of administered solution reaching nasopharyngeal walls. See the last visual under results for a pictorial representation of the 3D-printed soft nose used in the experiments.

## 3. Results

### 3.1. Optimal direction of spray axis and droplet sizes for effective targeting

Airflow and droplet tracking was simulated for spray nozzle placement in the left and right nasal airways of Subjects 1 and 2 under two standard inhalation rates (15 and 30 L/min), for drug droplet diameters 1 – 24 *μ*m, and for the two spray directions as per the CU and IU protocols. In all eight cases, the IU direction of the spray axis results in higher deposition at the nasopharynx in comparison to the CU protocol over a defined range of particle sizes (see Fig. 6). For instance: if we examine the deposition trends for spray administration through the right nostril of Subject 2 for the laminar regime inhalation (i.e. at 15 L/min), the peak nasopharyngeal deposition for IU is 46.5% for 13 *μ*m drug droplets (Fig. 6(b)), while the peak deposition for CU is only 0.53% for 14 *μ*m drug droplets (see again Fig. 6(b) and the corresponding zoomed-in for the CU delivery trends visual in Fig. 6(k)). In general, the droplet size range of ∼ 6 – 14 *μ*m is found conducive to targeted nasopharyngeal delivery with the IU protocol, considering a 2% cut-off for deposition efficiency of the tracked monodispersed droplet cluster of each size. The nearly hundred-fold increase in targeted deposition is remarkable and is achievable simply by re-orienting the spray axis from CU to IU.

**Figure 6:**
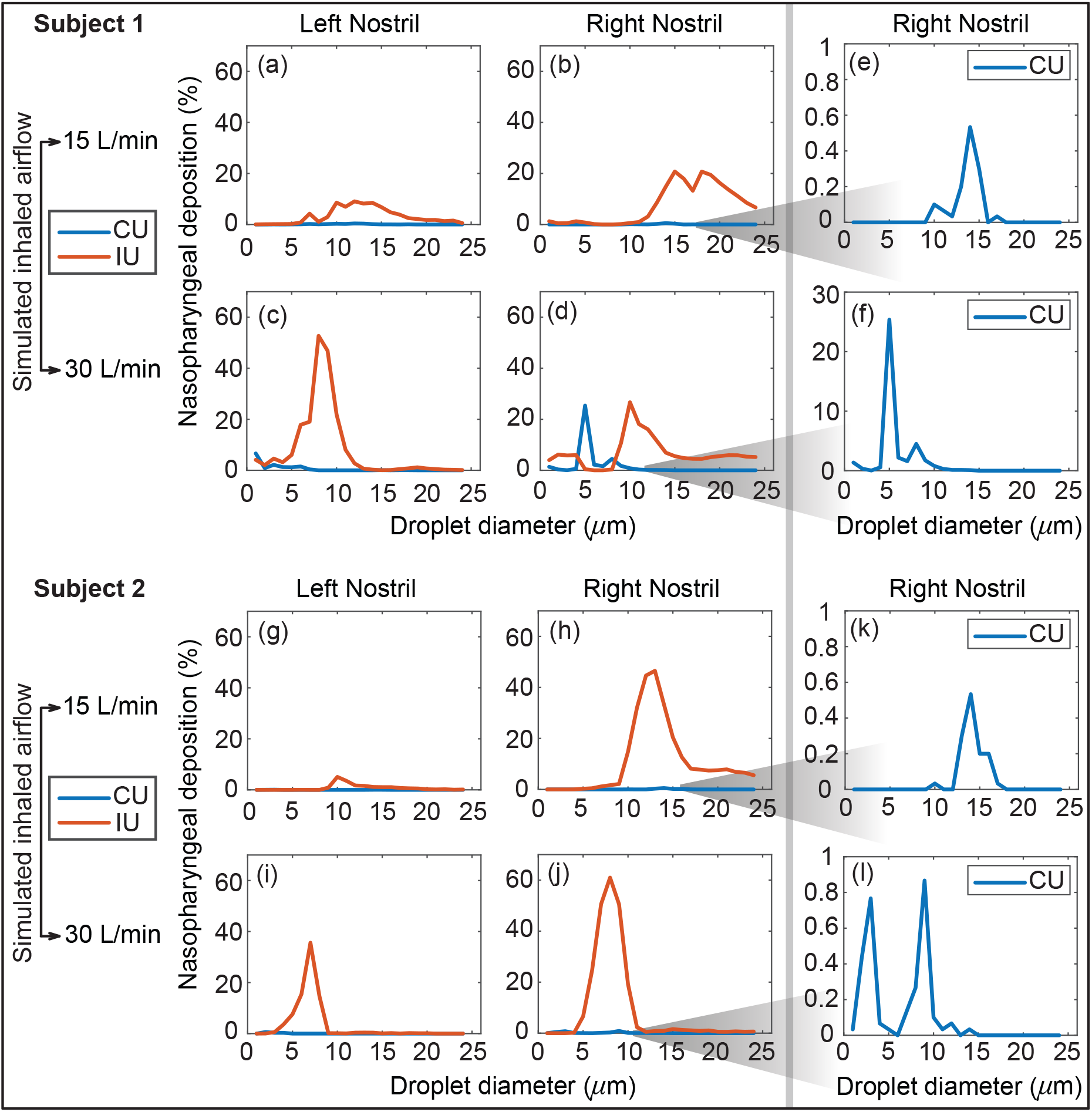
Panels (a) – (d) show the comparison of the regional deposition trends at the nasopharynx of Subject 1, for the IU and CU protocols, with monodispersed conical injections. The rows (a) – (b) are for 15 L/min inhalation; rows (c) – (d) are for 30 L/min inhalation. Panels (e) – (f) depict the representative zoomed-in trends for nasopharyngeal deposition with the CU protocol, on administering the spray through the right nostril. Similarly, panels (g) – (j) show the comparison of the regional deposition trends at the nasopharynx of Subject 2, for the IU and CU protocols. The rows (g) – (h) are for 15 L/min inhalation; rows (i) – (j) are for 30 L/min inhalation. Panels (k) – (l) depict the representative zoomed-in trends for nasopharyngeal deposition with the CU protocol, on administering the spray through the right nostril. The IU trend lines are marked in red; the CU trend lines are in blue. The reader should note the abbreviated vertical range on the (e) – (f) and (k) – (l) plots, prompted by the 2 orders-of-magnitude smaller deposition efficiency with CU.

### 3.2. Assessing sensitivity to IU perturbations

The variation of the nasopharyngeal deposition percentages over the assessed droplet size range (1 – 24 *μ*m) was compared between that of the IU protocol and for each of the PD 1 – 5 cases. Pearson’s correlation coefficient was greater than 0.5 for nearly every such comparison (see Fig. 7 and Table 1), showing a high degree of linearity between the new perturbed directions and the IU protocol. Moreover, the p-value associated with each correlation was much lower than the significance level, i.e. 0.05. This indicates that there is a statistically significant correlation between the simulation results on the targeted nasopharyngeal drug delivery for the IU and the perturbed directions. Physically, the satisfactory correlation between IU and PD 1 – 5 establishes the robustness of the IU spray protocol to user subjectivities.

**Table 1:** Statistical testing on the correlation between the regional deposition trends (for different drug droplet sizes) at the nasopharynx for the perturbed spray directions (i.e. PD 1 – 5), when compared to the nasopharyngeal deposition with the IU protocol.

| Model | Simulation Case | Pearson's Correlation Coefficient (r) |  |  |  |  | p-value associated with correlation |  |  |  |  |
| --- | --- | --- | --- | --- | --- | --- | --- | --- | --- | --- | --- |
|  |  | PD 1 | PD 2 | PD 3 | PD 4 | PD 5 | PD 1 | PD 2 | PD 3 | PD 4 | PD 5 |
| Subject 1 | Left Nostril Administration 15 L/min Inhalation | 0.6805 | 0.6745 | 0.9176 | 0.6155 | 0.6620 | 2.53E-04 | 3.01E-04 | 2.78E-10 | 1.37E-03 | 4.26E-04 |
|  | Right Nostril Administration 15 L/min Inhalation | 0.9526 | 0.8795 | 0.8469 | 0.8769 | 0.9580 | 7.43E-13 | 1.52E-08 | 1.80E-07 | 1.89E-08 | 2.01E-13 |
|  | Left Nostril Administration 30 L/min Inhalation | 0.8191 | 0.6122 | 0.9725 | 0.5525 | 0.9577 | 9.87E-07 | 1.48E-03 | 2.07E-15 | 5.12E-03 | 2.22E-13 |
|  | Right Nostril Administration 30 L/min Inhalation | 0.4607 | 0.7348 | 0.8711 | 0.5499 | 0.8029 | 2.35E-02 | 4.33E-05 | 3.06E-08 | 5.37E-03 | 2.34E-06 |
| Subject 2 | Left Nostril Administration 15 L/min Inhalation | 0.9548 | 0.6513 | 0.7114 | 0.7296 | 0.9343 | 4.49E-13 | 5.66E-04 | 9.72E-05 | 5.21E-05 | 2.49E-11 |
|  | Right Nostril Administration 15 L/min Inhalation | 0.9848 | 0.9629 | 0.9523 | 0.9805 | 0.9939 | 3.24E-18 | 5.29E-14 | 8.03E-13 | 4.77E-17 | 1.50E-22 |
|  | Left Nostril Administration 30 L/min Inhalation | 0.9348 | 0.8904 | 0.5591 | 0.7557 | 0.8534 | 2.29E-11 | 5.66E-09 | 4.51E-03 | 1.95E-05 | 1.16E-07 |
|  | Right Nostril Administration 30 L/min Inhalation | 0.9413 | 0.9512 | 0.9387 | 0.9877 | 0.9797 | 7.53E-12 | 1.02E-12 | 1.18E-11 | 3.05E-19 | 7.44E-17 |

**Figure 7:**
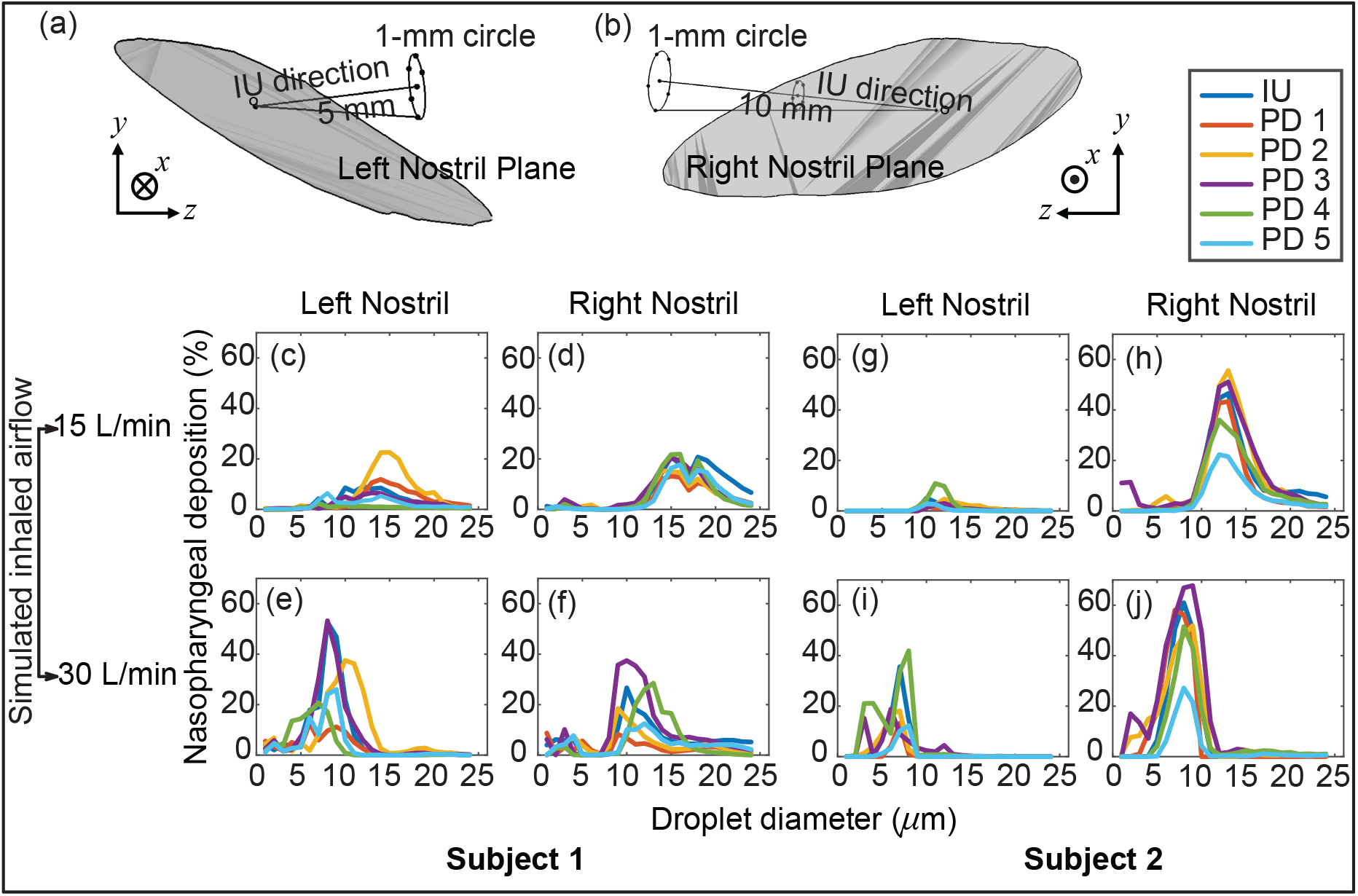
Panels (a) and (b) illustrate the *in silico* detection of the perturbed spray directions (PD), deviating slightly from the IU axis. The direction vectors are from the centroid of the nostril plane to the points lying on a 1-mm circle that is 5 mm and 10 mm (respectively for the left and right nostril placement) from the nostril plane centroid (see Section 2.4 for associated details). Panels (c) – (f) for Subject 1 and panels (g) – (j) for Subject 2 compare the respective nasopharyngeal deposition trends for PD 1 – 5 directions, with respect to that of the “Improved Use” (IU) protocol. The top row is for 15 L/min inhalation; the bottom row is for 30 L/min inhalation rate. Clustering of the plots signifies robustness of the IU usage parameters; in other words, the IU protocol is satisfactorily less sensitive to user subjectivities.

### 3.3. Verification of optimal droplet sizes through scaling analysis

The droplet size ranges that registered peak nasopharyngeal deposition under each inhalation condition were further analyzed and validated for reliability, through a Stokes number-based scaling analysis.^39^ The Stokes number (St) is defined as^32^

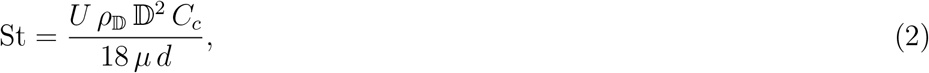

where *U* for the present system is the airflow rate divided by flux area, *ρ*_𝔻_ is the material density of the inhaled droplets, *C*_*c*_ is the Cunningham slip correction factor, *μ* is the dynamic viscosity of the ambient medium i.e. air, and *d* represents the characteristic diameter of the flux cross-section. Now, all other flow and morphological parameters staying invariant, Equation 2 directly leads to the following scaling law:

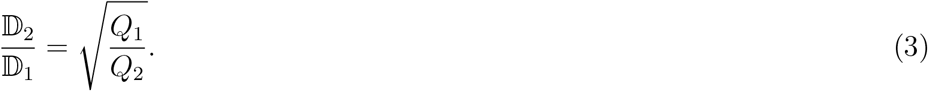

Herein (*Q*_*i*_, 𝔻_*i*_) are different airflow rate and droplet size pairings. Let us now consider a representative example, say the right nostril spray administration in Subject 2. For at least 2% nasopharyngeal deposition, the computationally predicted ideal droplet size range during 30 L/min inhalation is [𝔻min, 𝔻max] = [5, 11] *μ*m. Equation 3 can consequently help us to project the corresponding ideal size range at the lower inhalation rate of 15 L/min. If the to-be-projected droplet size range that would generate peak nasopharyngeal deposition during the latter case is represented by 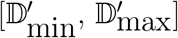 in *μ*m, then

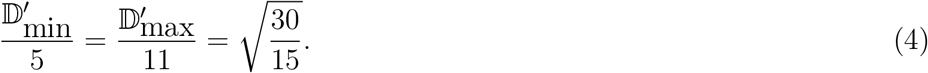

This results in 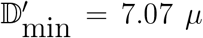 and 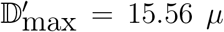. Despite the simplicity of this scaling analysis, the computationally identified range 9 – 24 *μ*m for the same breathing conditions hence follows the same trend on the number scale, in terms of the respective variations from the extremal limits of [𝔻min, 𝔻max]. The penultimate panel in Fig. 8 visually illustrates this specific example; see the remaining panels in Fig. 8 for all the other test cases. The directional change of the extremal limits for the St-projected ideal droplet size ranges remarkably agrees with the corresponding CFD-based size ranges in all cases, except in one trivial outlier: see panel (c) for Subject 1’s right nostril, there the maximum ideal size limits for both 15 and 30 L/min are 24 *μ*m; the St-projected maximum ideal droplet size for 30 L/min is, however, 33.94 *μ*m.

**Figure 8:**
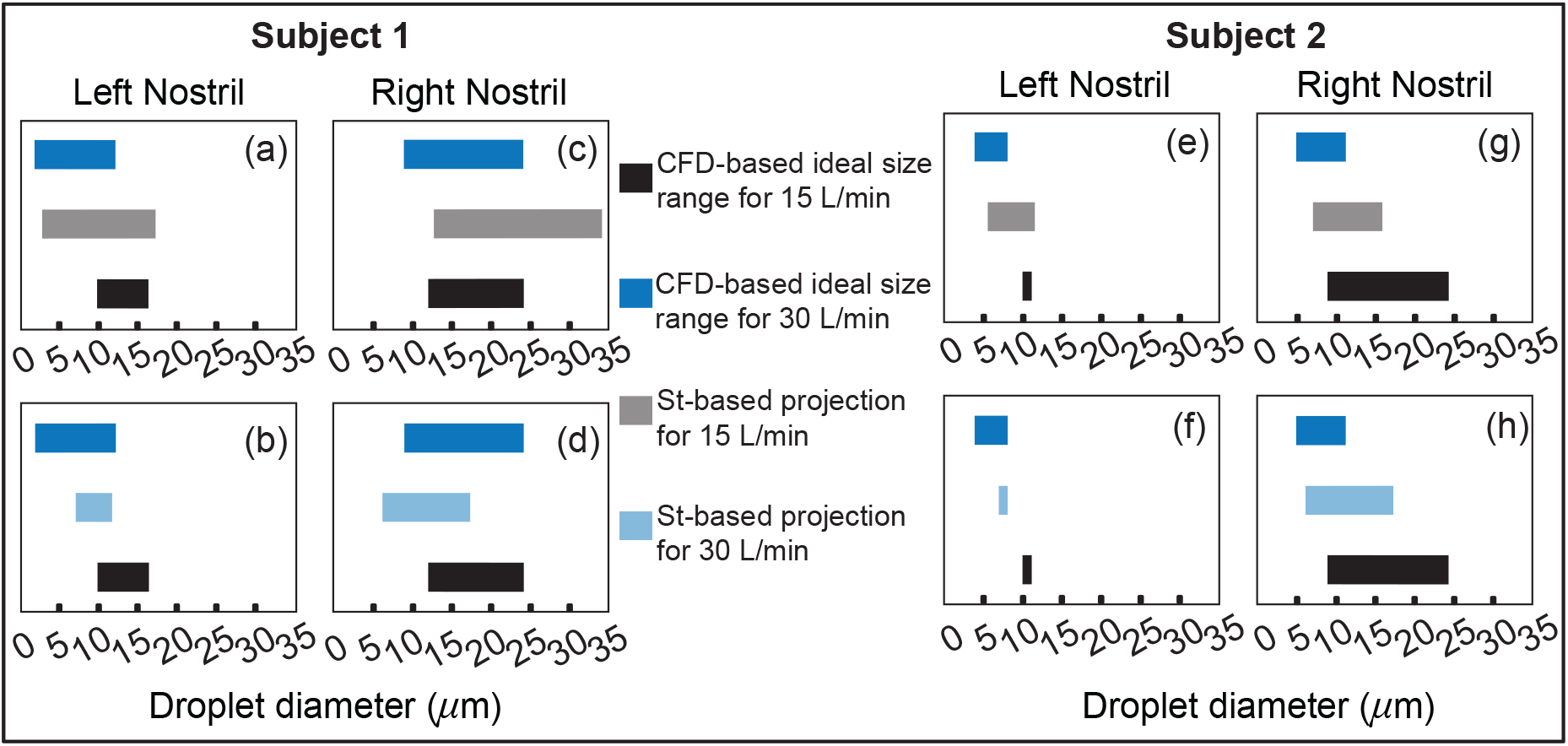
Panels (a) – (d) for Subject 1 and panels (e) – (h) for Subject 2 visually depict the Stokes number (St)-based projections of ideal droplet size ranges for maximal targeted deposition at the nasopharynx. The directional change of the St-projected ranges agree with the corresponding CFD-based ideal droplet size ranges in all the test cases, except in one trivial outlier: see panel (c), where the maximum ideal size limits for both 15 and 30 L/min are 24 *μ*m; the St-projected maximum ideal droplet size for 30 L/min is, however, 33.94 *μ*m. See Section 3.3 for a representative discussion for the data reported in (g).

### 3.4. Comparison of the in silico findings to physical experiments

Panel (a) in Fig. 9 portrays the order-of-magnitude improvement in targeted drug deposition at the nasopharynx (with the IU protocol over the CU protocol), when taking into the account the droplet size distributions^15, 18^ in real over-the-counter spray products, *viz*. Flonase™ and Nasacort™, in an administered shot. See Section 2.5 for the related study methods. Considering all the test cases, the average IU-over-CU improvement for the two chosen spray products, as projected from the CFD simulations, was 2.117 orders-of-magnitude with a standard deviation of 0.506 orders. The physical experiments in Subject 3 (presenting an entirely different anatomy) show a comparable improvement in nasopharyngeal delivery, by 2.215 orders-of-magnitude, with a standard deviation of 0.016 orders. Panel (b) in Fig. 9 plots the experimental measurements. Hence, the computational predictions differ from the *in vitro* data by less than 5%, thereby lending robust support to the implemented *in silico* framework.

**Figure 9:**
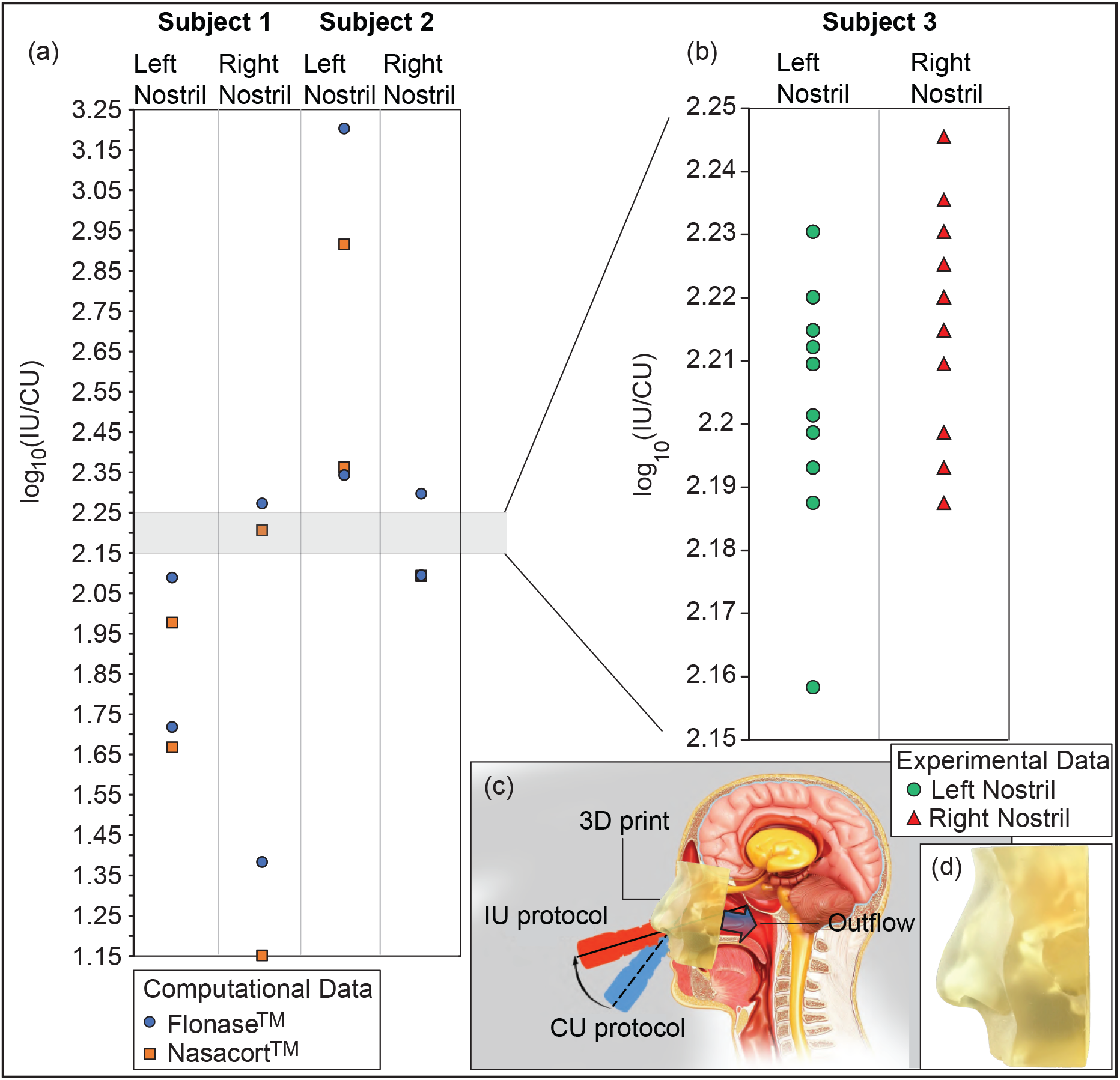
Panel (a) shows the order-of-magnitude IU-induced improvement in drug mass deposits at the nasopharynx of Subjects 1 and 2 (when compared to the CU delivery numbers), while considering the droplet size distribution in each administered shot of two common over-the-counter spray products: Flonase™ and Nasacort™. Panel (b) shows the measurements from a set of physical experiments with sprayed watery solution in different Subject 3. As an indicator for agreement between the computational and experimental projections, the vertical range in (b) is a medial subset of that in (a). Note that several data-points roughly superimposed over each other, in both (a) and (b). Panel (c) presents a cartoon of the experimental setup. A separate in-set visual for the 3D-printed soft nose, with realistically pliable external nares, is shown in (d).

## 4. Discussion

- *On inputs to targeted drug design –* With targeted delivery of pharmaceutic agents to the viral infection hot-spots in the posterior upper airway (e.g. at the nasopharynx) a clear challenge,^15, 40, 41^ the experimentally-validated findings (see Fig. 6) from this study point to the droplet size range of ∼ 6 – 14 *μ*m as being the most effective at maximizing the sprayed and inhaled percentage deposition at the clinical upper airway target site for SARS-like infections. The information could be utilized to design next-generation intranasal drug formulations, along with novel spray devices and atomizers.
- *On inputs for effective spray usage strategies –* The significant 2 orders-of-magnitude improvement (see Fig. 9) in nasopharyngeal delivery of intranasally sprayed drugs with the new IU protocol, over the typically recommended CU protocol, clearly warrants a revisit of the standard usage instructions for existing nasal spray products. While Section 2.3 lays out the different criteria points for *in silico* detection of the IU direction †; in ordinary language: the user can replicate the IU protocol by holding spay nozzle as horizontally as possible at the nostril, with a slight tilt towards the cheeks and pointed a little at the outer edge of the eye (e.g., right eye if one is placing the spray at the right nostril). See Fig. 10 for a sample demonstration.

**Figure 10:**
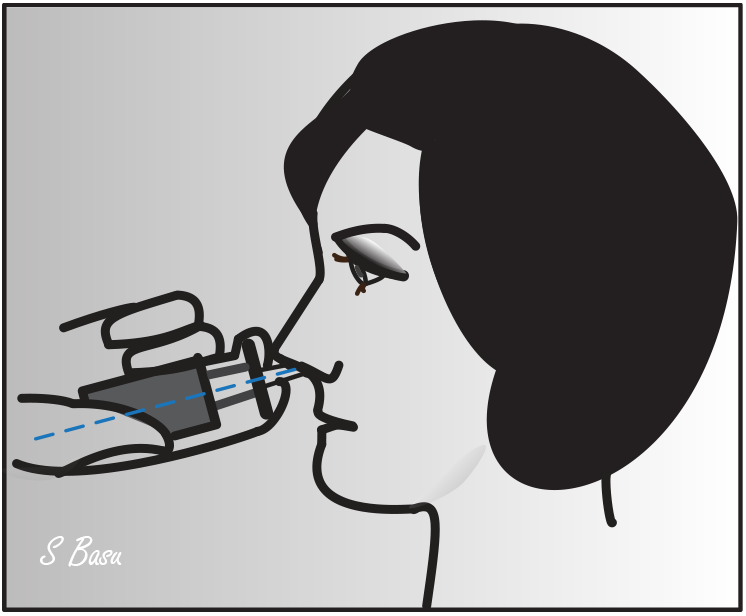
Demonstrative sagittal sketch for the “Improved Use” (IU) protocol, outlining how to hold a spray bottle during intranasal administration. Illustration is by the lead author.
- *On the limitations of respiratory flow modeling –* The reader should note that a realistic modeling of mucociliary transport along the morphologically complex airway cavity constitutes a significant open question in the domains respiratory transport mechanics.^42^ In this study, we have implemented state-of-the-art algorithms to identify the droplet sizes that are efficient at *direct* nasopharyngeal delivery, under the impact of inhaled airflow when sprayed into the intranasal space. However, a big caveat lies in what happens to the larger droplets that happen to deposit along the anterior parts of the airway. Quantifying their mucus-driven downstream transport mechanics and correlating that with the therapeutic efficacy of the drug solutes when they reach the posterior clinical target sites poses a major translational challenge, to be addressed by the community in future.
- *On the constraints posed by the reconstructed in silico geometries –* The CT-based anatomically realistic reconstructions, while accurately replicating the topological convolutions implicit in a real tortuous respiratory cavity, still come with the caveat of containing structurally rigid airway walls. However, though the rigidity of the walls (intended to mimic the internal tissue surfaces and cartilages) is somewhat unrealistic, the time-scale of inhaled transport is on the scale of 10^−1^ s.^18^ and the idealization could be considered a mechanically feasible assumption that is sufficient to extract the fundamental nuances underlying such physiologically complex transport processes.
- *On the usability of the findings despite the small test cohort –* Clearly the current study is somewhat limited given the restricted sample size of only two main test subjects (i.e. Subjects 1 and 2). However, the congruity in targeted delivery improvement (see Section 3.4 and Fig. 9) in a randomly-selected third subject (Subject 3) bodes well for the general extensibility of the essential findings to a wider cohort. At the least, the results presented here, though preliminary in essence given the small cohort size, could be considered an important step in the mechanistic characterization of the respiratory transmission dynamics for improving the performance of intranasally administered spray products.
- *On toxicity evaluation –* Any new formulation or drug delivery device that might attempt to replicate the improved targeted deposition at intranasal sites, based on the current findings, will essentially form a surface contacting mechanism with limited duration contact. For determination of usage safety levels, such a development will also require biocompatibility testing of the device, including check of three basic biocompatibility endpoints (i.e., cytotoxicity, irritation,^43^ sensitization) per the Food and Drug Administration (FDA) guidance,^44, 45^ by providing test data and/or relevant justification (e.g., history of clinical use for the same device).

## 5. Final takeaways

Intranasal sprays could represent a useful administration strategy for nasal hygiene products, prophylactics, and antivirals – for respiratory pathogens that would first trigger an upper airway infection, such as SARS-CoV-2. In this study, we have used computational fluid dynamics simulations to illustrate that simple tweaks to the nasal spray direction can result in vastly improved drug delivery to the critical viral infection sites inside the nose, more specifically the delivered dose registers an approximately 2 orders-of-magnitude improvement. The proposed IU protocol (see Figs. 3 and 4; also Fig. 10) is easy-to-replicate and has been verified to be robust to small perturbations that may stem from user subjectivities. Also, the droplet size range of ∼ 6 – 14 *μ*m is found most efficient at facilitating direct delivery of intranasally sprayed drug particulates at the nasopharynx, which is the dominant infection trigger zone. Both these key pieces of findings bear the promise for developing increasingly effective intranasal pharmaceutic formulations, along with upgraded designs for nasal drug delivery devices and atomizers.

## Data Availability

All data produced in the present study are available upon reasonable request to the corresponding author.

## Acknowledgements

The authors thank the *Bioinspiration & Biomimetics* editorial board for the invitation to publish. This material is based on work partially supported by the National Science Foundation (NSF) RAPID Grant 2028069 for COVID-19 research, with SB as the Co-Principal Investigator. Any opinions, findings, and conclusions or recommendations expressed here are, however, those of the authors and do not necessarily reflect views of the NSF.

## Author contributions

SB, DJM, AC conceived this study; SB developed the study protocol, the anatomic reconstructions and drafted the manuscript; MMHA carried out the physical experiments and the theoretical analysis; SB, YL, PAB, PA, NKK performed the numerical simulations; AM, ZS processed the computational data; JS tested the over-the-counter spray products; SB and DJM jointly supervised the student researchers (MMHA, YL, PAB, PA, NKK, AM, ZS).

## Footnotes

† (i) Extended spray axis for the IU protocol intersects the nasopharynx; (ii) as a condition for clinical safety (based on recommendation from attending rhinologists^15, 37^), the axis must not cut through the septum; (iii) the spray axis should intersect the lateral wall of the nasal cavity as posteriorly as feasible.

